# COVID-19 test positivity: predictive value of various symptoms in a large community-based testing program in California

**DOI:** 10.1101/2021.03.03.21252014

**Authors:** Dave P. Miller, Scott Morrow, Robert M. Califf, Cameron Kaiser, Ritu Kapur, Casimir Starsiak, Jessica Mega, William J. Marks

## Abstract

**Background:** Much of the early data on COVID-19 symptomatology was captured in the hospital setting. In a community setting the symptoms most predictive of SARS-CoV-2 positivity may be different. Data from the California sites of a COVID-19 community testing program are presented here.

**Methods:** Prior to being tested, participants in the Baseline COVID-19 Testing Program completed an online screener, in which they self-reported basic demographics and the presence or absence of 10 symptoms. Both positive and negative COVID-19 RT-PCR tests were linked back to the screener data. A multivariable model of positivity was fit using generalized estimating equations, adjusting for month of testing as a fixed effect and accounting for clustering of data within each test site.

**Results:** Among 547,018 first-time tests in California in 2020, positivity rates were 3.4%, 9.9%, and 19.8% for participants with no symptoms, 1 symptom, or 2 or more symptoms at the time of screening, respectively. All ten symptoms were individually associated with higher positivity rates, but only six of ten symptoms were associated with higher positivity when adjusting for other symptoms. Major symptoms with highest predictive value were recent loss of taste or smell, fever, and coughing with ORs of 3.27, 1.97, and 1.95, respectively. Shortness of breath and vomiting or diarrhea were negatively associated with positivity adjusting for other symptoms and, absent other symptoms, participants with these symptoms did not have significantly higher positivity rates than asymptomatic participants.

**Conclusions:** Recent loss of taste and smell should be elevated to a major symptom along with fever and coughing in public health messaging and in our community approach to testing and surveillance, while mild to moderate shortness of breath should be de-emphasized as a sensitive early predictor of COVID-19 positivity.

## Introduction

In late 2019, the identification of a novel coronavirus in the city of Wuhan, China preceded a country-wide outbreak that quickly escalated into a pandemic by March of 2020 [1]. Early studies of the clinical course of COVID-19, the disease caused by SARS-CoV-2, most commonly reported symptoms of fever, cough, shortness of breath, and myalgia [2,3,4]. Other commonly reported symptoms included headache, fatigue, congestion, nausea, vomiting, and diarrhea. These studies, published during the early stages of the pandemic, were largely based on populations of hospitalized people who were critically ill. It is possible that symptoms vary over the clinical course of the disease, and that the symptoms present at the time of hospitalization are not those most useful for screening and management of an ambulatory population.

As the course of the pandemic has progressed, the focus of public health officials has expanded from the acute needs of critically ill people to the population at large. Symptom lists are used by public health officials to advise the population on when to get tested or consider self-quarantine, and to advise businesses on which customers should not be allowed entry. For example, though it was rarely reported in studies of hospitalized individuals, it has been recently hypothesized that acute loss of taste or smell has predictive value for higher likelihood of SARS-CoV-2 infection [5,6], that it should be considered sufficient to justify testing and self-isolation, [7] and that it might indicate a milder course of disease [6].

There are additional challenges posed by the known asymptomatic / presymptomatic transmission of the disease [8,9]. A better understanding of the odds of being infected with SARS-CoV-2 given a specific set of symptoms, and how this compares to those who do not currently have symptoms, could better arm public health officials and the public as they make decisions about mitigation measures and individual behavior.

Here we present data from our California sites from the Baseline COVID-19 Testing Program, focusing on the aspects of the data that we believe are most relevant for developing actionable public health recommendations as the pandemic continues.

## Subjects, Materials, Methods

### Testing Program

The Baseline COVID-19 Testing program was launched in March of 2020. Participants completed an online screener survey and were next allowed to schedule a test at a community testing site. The screener included the language “this program is intended to expand access to COVID-19 risk screening and testing. It is not intended for people experiencing severe symptoms such as severe cough, severe shortness of breath, severe fever, or other concerning symptoms who may need more immediate medical care.” Beyond discouraging the participation of those with severe symptoms, program inclusion and testing criteria went through several iterations prior to May of 2020, at which time the core content of the current version of the screener was solidified. Participants consented to the use of the data for public health purposes, which has included reporting on summary data and unique program insights to California county public health officials.

### Testing Sites

In 2020, the program had over 400 community testing sites in 16 states, including 195 California testing sites. Among the California sites, 113 were in Rite Aid stores (through its partnership with the U.S. Department of Health and Human Services) and 82 were operated by Verily Life Sciences for the California Department of Public Health, 6 of which were later operated by Verily for San Mateo County. At least one site was present in 48 of California’s 58 counties. All tests were performed using RT-PCR at labs operated by Quest, Bioreference Laboratory, LabCorp, Eurofins, or Verily.

### Screener

Prior to scheduling an appointment, all screener questions had to have been completed online. The self-reported screener data includes basic demographics, contact with a confirmed case, occupational risk, 10 symptoms, and comorbidities. After the lab completes the test, the screener data is linked to the test results for both positive and negative tests.

### Statistical Methods

Analyses were conducted for participants with complete data, excluding indeterminate tests and tests that preceded the introduction of the current core screener questions. Test positivity was the binary outcome used in a series of generalized estimating equation models. The models use a logit link, similar to logistic regression, but take into account within-site clustering, which is expected due to the geographic nature of the pandemic. Additionally, both the single symptom and multi-symptom models adjust for month of test using fixed effects. Models were fit using the statsmodels gee package for python. Confidence intervals were computed using the Wald method.

## Analysis & Results

### Cohort

Over 2 million tests were performed in 2020, including 916,177 in California. Among those, 758,497 had complete screener data. After excluding re-tests and indeterminate tests, more than half a million tests (547,018) were available for analysis (Figure 1).

**Figure 1:**
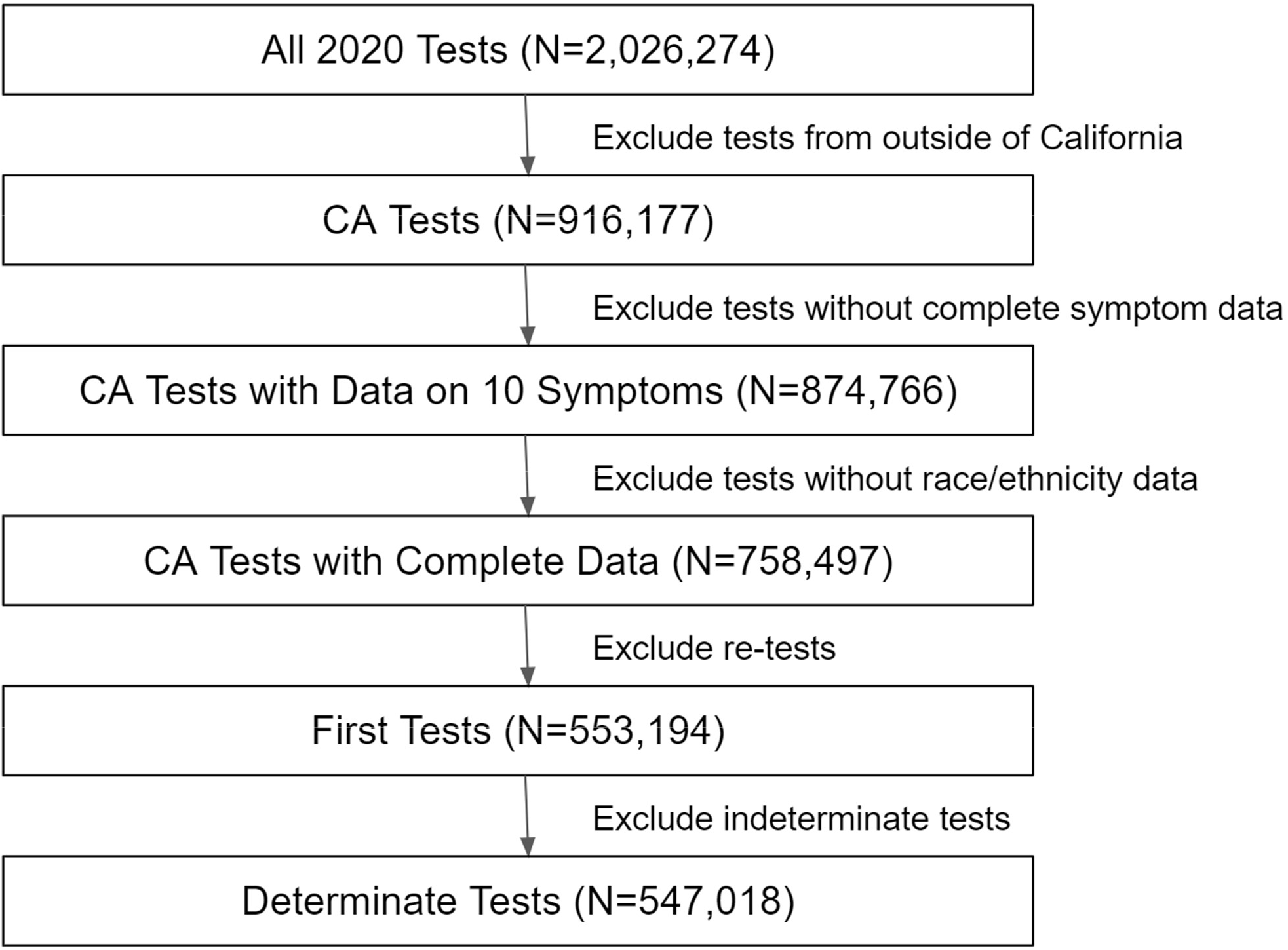
STROBE diagram

### Demographics, Symptoms, and Positivity

More than half of the participants (318,176) were asymptomatic at the time of screening (Table 1a). An additional 73,886 had only one single symptom out of the list of 10 symptoms presented on the screener, and 154,956 had two or more self-reported symptoms. More females than males were tested in all 3 groups, and the majority were 18-44 years old. Participants 65 and older represented 9.2% of the asymptomatic cohort vs 4.9% of the cohort with multiple symptoms. Hispanics were a smaller proportion of the asymptomatic cohort vs the multi-symptom cohort (30.1% vs 45.5%), while the opposite was true of Asians (24.2% vs 14.9%). Whites were a minority in all cohorts, reflecting California’s demographics.

**Table 1a:** Demographics

|  |  | Symptoms at Time of Screening |  |  |
| --- | --- | --- | --- | --- |
|  |  | Asymptomatic | Single Symptom | Multiple Symptoms |
| Total | All | 318,176 (100.0%) | 73,886 (100.0%) | 154,956 (100.0%) |
| Gender | Female | 170,169 (53.5%) | 41,671 (56.4%) | 89,327 (57.6%) |
|  | Male | 147,718 (46.4%) | 32,142 (43.5%) | 65,443 (42.2%) |
| Age | 13-17 | 2,319 (0.7%) | 548 (0.7%) | 1,235 (0.8%) |
|  | 18-24 | 60,205 (18.9%) | 15,067 (20.4%) | 33,752 (21.8%) |
|  | 25-34 | 93,797 (29.5%) | 24,106 (32.6%) | 49,508 (31.9%) |
|  | 35-44 | 51,014 (16.0%) | 13,012 (17.6%) | 27,940 (18.0%) |
|  | 45-54 | 41,759 (13.1%) | 9,135 (12.4%) | 20,165 (13.0%) |
|  | 55-64 | 39,763 (12.5%) | 7,360 (10.0%) | 14,712 (9.5%) |
|  | 65-74 | 22,220 (7.0%) | 3,577 (4.8%) | 5,948 (3.8%) |
|  | 75+ | 7,098 (2.2%) | 1,081 (1.5%) | 1,696 (1.1%) |
| Ethnicity | Hispanic | 95,922 (30.1%) | 28,597 (38.7%) | 70,568 (45.5%) |
|  | Non-hispanic | 222,254 (69.9%) | 45,289 (61.3%) | 84,388 (54.5%) |
| Race | Asian | 76,860 (24.2%) | 15,519 (21.0%) | 23,141 (14.9%) |
|  | Black or African American | 17,003 (5.3%) | 4,000 (5.4%) | 7,500 (4.8%) |
|  | Hawaiian or Pacific Islander | 6,266 (2.0%) | 1,310 (1.8%) | 2,656 (1.7%) |
|  | Native American | 5,321 (1.7%) | 1,492 (2.0%) | 3,589 (2.3%) |
|  | White | 156,154 (49.1%) | 33,768 (45.7%) | 74,431 (48.0%) |

\* Percentages do not add to 100 for race because participants could select “Other” and because participants could select more than one race.

Positivity rates were dramatically higher in symptomatic participants vs asymptomatic participants for every demographic group (Table 1b) and similarly much higher with multiple symptoms versus a single symptom. Among Hispanics, one of the most disproportionately affected cohorts, positivity rates were 6.6%, 15.7%, and 28.1% with 0, 1, or 2+ symptoms, respectively. Among Asians, who have had some of the lower rates in California, positivity rates were 1.8%, 5.5%, and 11.9% with 0, 1, or 2+ symptoms, respectively.

**Table 1b:** Positivity rate by demographics and symptom status

|  |  | Symptoms at Time of Screening |  |  |
| --- | --- | --- | --- | --- |
|  |  | Asymptomatic | Single Symptom | Multiple Symptoms |
| Total | All | 3.4% | 9.9% | 19.8% |
| Gender | Female | 3.3% | 9.0% | 18.4% |
|  | Male | 3.6% | 11.0% | 21.7% |
| Age | 13-17 | 13.0% | 31.2% | 41.9% |
|  | 18-24 | 4.2% | 11.2% | 20.3% |
|  | 25-34 | 3.0% | 8.4% | 17.6% |
|  | 35-44 | 3.2% | 9.0% | 19.4% |
|  | 45-54 | 3.7% | 10.9% | 22.2% |
|  | 55-64 | 3.0% | 10.4% | 21.0% |
|  | 65-74 | 2.8% | 10.1% | 19.9% |
|  | 75+ | 3.6% | 12.6% | 23.2% |
| Ethnicity | Hispanic | 6.6% | 15.7% | 28.1% |
|  | Non-hispanic | 2.1% | 6.2% | 12.9% |
| Race | Asian | 1.8% | 5.5% | 11.9% |
|  | Black or African American | 3.1% | 9.0% | 15.8% |
|  | Hawaiian or Pacific Islander | 2.9% | 8.1% | 17.5% |
|  | Native American | 4.4% | 10.0% | 18.4% |
|  | White | 2.7% | 7.9% | 16.1% |

### Temporal trends

Positivity rates in California had a peak in July, followed by steady improvement through October, followed by a sharp increase in positivity rates in November and December (Figure 2). Positivity rates for asymptomatic participants followed the same trend as for those with symptoms. The December outbreak has been so acute that the positivity rate among asymptomatic participants at the end of the year exceeded the late Spring positivity rate for participants with multiple symptoms.

**Figure 2:**
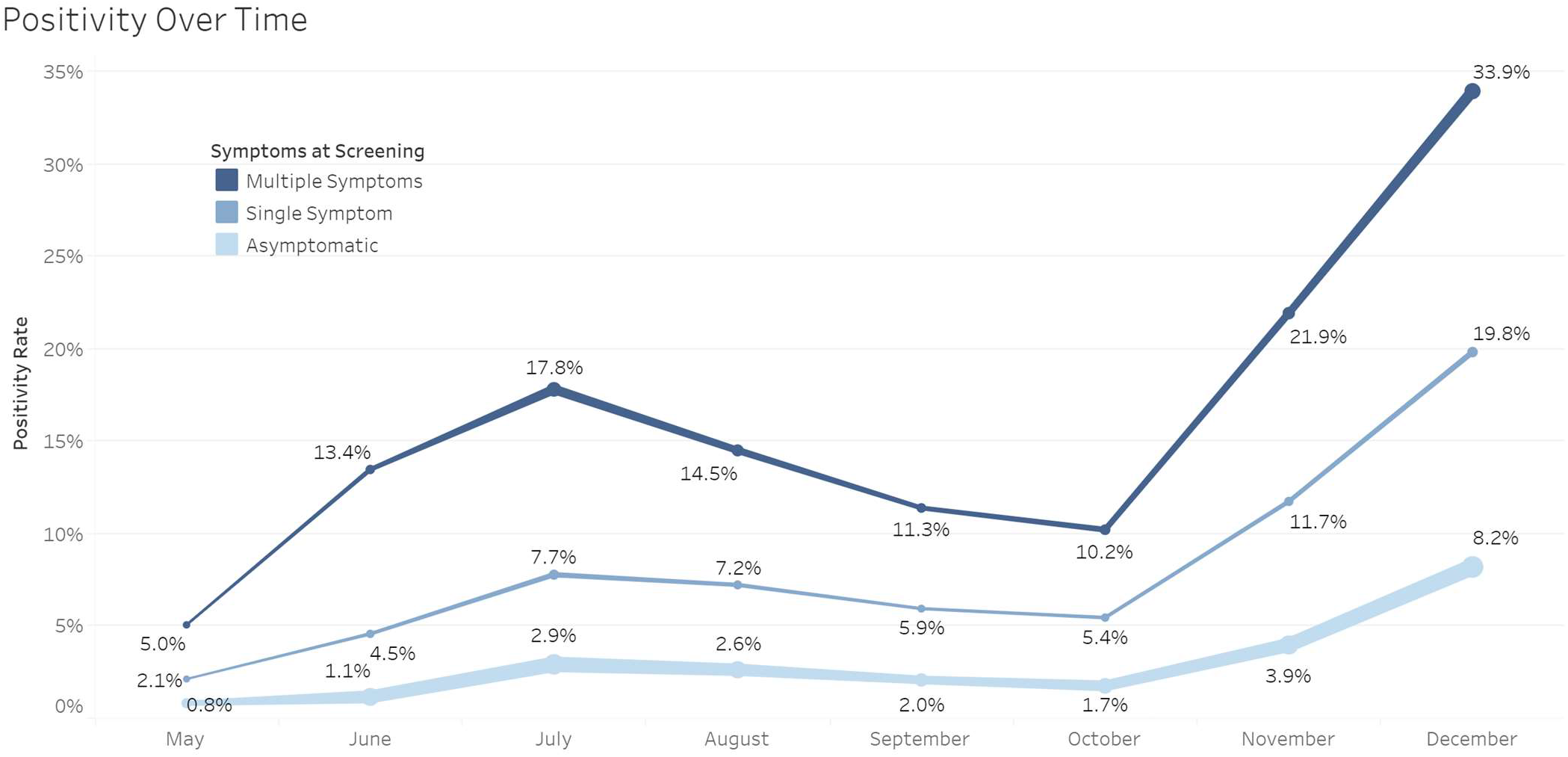
Positivity over time Note: line width indicates test volume.

### Symptom prevalence

The four most common symptoms reported on the screener were coughing, headache, muscle pain, and sore throat, and these same four symptoms were the most common among positive tests and also among negative tests. Coughing (47.2%) and headache (46.0%) were the most frequently reported symptoms among those with a positive test, while headache (19.9%) and sore throat (17.6%) were the most frequently reported among those with negative tests. Every individual symptom was more common among participants with positive tests than those with negative tests.

### Symptom prognostic value

Single symptom models (Figure 3) show that every symptom is individually associated with positive tests, with odds ratios ranging from 5.75 for recent loss of taste and smell (95% CI 5.47-6.06) to 1.52 for shortness of breath (95% CI 1.46-1.57). A multivariable model, including all 10 symptoms, shows that only 6 of the symptoms are associated with positive tests, conditional on the presence or absence of the other symptoms.

**Figure 3:**
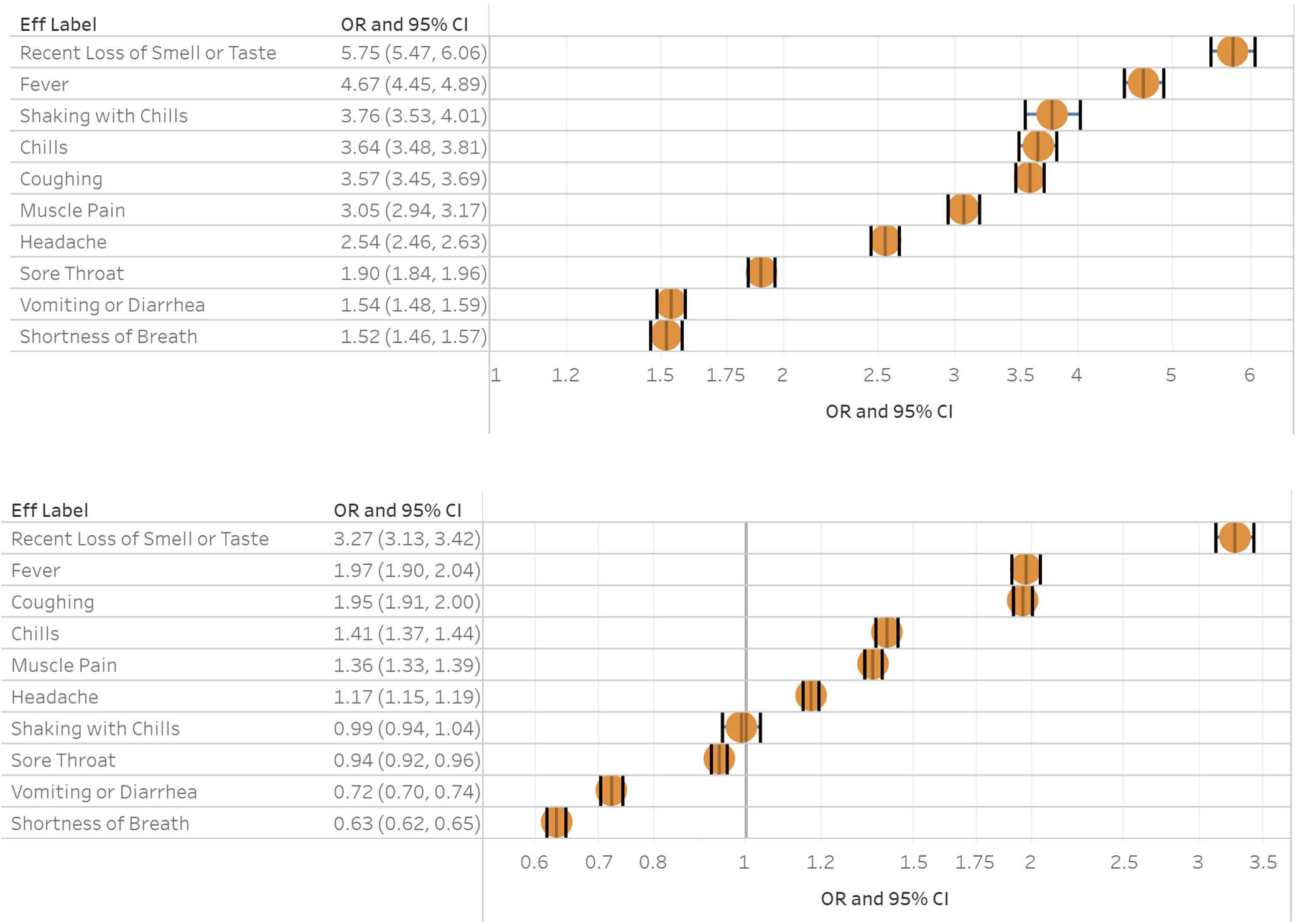
Odds Ratios for each of 10 symptoms, without adjustment for other symptoms and with adjustment for other symptoms

Recent loss of taste or smell, fever, and coughing have multivariable ORs of 3.27, 1.97, and 1.95, respectively. For summary purposes, these will be referred to as major symptoms. The three symptoms with weaker, but still positive, associations are chills, muscle pain, and headache, with ORs of 1.41, 1.36, and 1.17, respectively. For summary purposes, these will be referred to as supplementary symptoms. After adjusting for other symptoms, the models shows that sore throat, vomiting or diarrhea, and shortness of breath are not associated with a higher chance of a positive test,.

### Proposed symptom hierarchy

Although the 3 major symptoms are not all of equal prognostic value and the 3 supplementary symptoms are not all of equal prognostic value, a simple summary can nonetheless be created by considering the count of each kind of symptom. Table 3 shows the way the positivity rate increases with each additional major symptom and each additional supplementary symptom, with and without the negatively associated symptoms.

**Table 3:** Positivity rates by count of major symptoms and count of supplementary symptoms among participants with and without SOB, patients with and without GI problems, and with and without sore throat)

| Supplementary Symptoms | Count of major symptoms when shortness of breath is **absent** |  |  |  | Count of major symptoms when shortness of breath is **present** |  |  |  |
| --- | --- | --- | --- | --- | --- | --- | --- | --- |
|  | 0 | 1 | 2 | 3 | 0 | 1 | 2 | 3 |
| 0 | 3.5% | 17.0% | 37.5% | 34.4% | 3.9% | 8.5% | 23.6% | 25.7% |
| 1 | 7.2% | 19.9% | 41.4% | 51.9% | 4.6% | 11.6% | 24.6% | 31.5% |
| 2 | 9.9% | 25.2% | 46.2% | 55.5% | 6.9% | 14.7% | 30.8% | 37.1% |
| 3 | 16.3% | 29.6% | 52.8% | 63.4% | 9.9% | 19.5% | 36.0% | 48.0% |

| Supplementary Symptoms | Count of major symptoms when GI symptoms are **absent** |  |  |  | Count of major symptoms when GI symptoms are **present** |  |  |  |
| --- | --- | --- | --- | --- | --- | --- | --- | --- |
|  | 0 | 1 | 2 | 3 | 0 | 1 | 2 | 3 |
| 0 | 3.5% | 16.0% | 35.1% | 31.1% | 4.1% | 12.8% | 29.4% | 32.0% |
| 1 | 7.2% | 19.0% | 38.6% | 44.7% | 5.1% | 13.1% | 30.4% | 44.8% |
| 2 | 10.2% | 24.2% | 44.7% | 50.4% | 6.1% | 16.0% | 30.9% | 43.1% |
| 3 | 17.0% | 30.3% | 51.4% | 59.2% | 9.9% | 18.6% | 36.4% | 49.5% |

| Supplementary Symptoms | Count of major symptoms when sore throat is **absent** |  |  |  | Count of major symptoms when sore throat is **present** |  |  |  |
| --- | --- | --- | --- | --- | --- | --- | --- | --- |
|  | 0 | 1 | 2 | 3 | 0 | 1 | 2 | 3 |
| 0 | 3.4% | 16.9% | 37.8% | 32.8% | 5.4% | 13.5% | 28.7% | 28.6% |
| 1 | 7.0% | 20.1% | 42.5% | 52.0% | 6.8% | 16.0% | 32.0% | 38.7% |
| 2 | 9.3% | 24.3% | 47.8% | 57.2% | 9.8% | 20.8% | 36.7% | 43.1% |
| 3 | 16.4% | 27.7% | 49.9% | 61.3% | 13.3% | 25.9% | 44.6% | 52.8% |

For example, the positivity rate with one major symptom and two supplementary symptoms is 25.2% in participants who do not report shortness of breath compared to 14.7% in participants who do report shortness of breath, 24.2% vs 16.0% for those who do not report vomiting or diarrhea vs those who do, and 24.3% vs 20.8% for those who do report sore throat vs those who do.

Adding one of the three non-predictive symptoms to one or more major or supplementary symptoms almost always results in a lower positivity rate; however, the pattern is slightly different for those participants who have no major or supplementary symptoms. For example, participants with no major or supplementary symptoms, the positivity rate is 3.4% for those who do not report a sore throat vs 5.4% for those who do. Participants whose sole symptom is either shortness of breath or gastrointestinal in nature (n=9,583) have a numerically higher positivity rate than asymptomatic participants (n=338,627), but the difference is not statistically significant (3.5% vs 3.9%, p=0.08). Among 375 participants whose only two symptoms are shortness of breath and GI-related, there is also a numerically higher non-significant difference (5.1% vs 3.5%, p=0.11).

Lastly, we assessed whether or not the non-predictive symptoms continued to be non-predictive during periods of high overall prevalence. In December, the positivity rates for participants with zero symptoms, one symptom, or more than one symptom rose to 8.2%, 19.8%, and 33.9% respectively. For those with exactly one symptom in December, if that symptom was one of the two non-predictive symptoms (shortness of breath or GI-related), the positivity rate was 8.3%, nearly identical to the positivity rate for asymptomatic participants. For those with exactly two symptoms, if those symptoms were the non-predictive symptoms of shortness of breath and GI-related, the positivity rate was 5.9%.

## Discussion

In the community testing setting, we determined 3 major symptoms: recent loss of taste or smell, fever, and coughing. Frontdoor screening to enter businesses should focus foremost on these three symptoms. Supplementary symptoms of headache, muscle pain, and chills are also prognostic and should be considered in conjunction with major symptoms when making a determination about the urgency of getting tested or a determination to self-quarantine while waiting for a test result.

Sore throat alone shows a modestly higher positivity rate, but its presence in conjunction with other symptoms decreases the positivity rate. It is not clear from these data what role sore throat should have in public health communications and screening activities. Shortness of breath and GI symptoms are not positively associated with a positive test, after adjustment for the major and supplementary symptoms. When present as single symptoms, positivity rates are not statistically different from positivity rates in asymptomatic participants. Persons with these symptoms, without any major or supplementary symptoms, should therefore not be considered at any higher risk than asymptomatic persons.

This analysis has several limitations. There is undoubtedly selection bias, both in the exclusion of symptomatic people who choose not to get tested and among people with severe symptoms, who are not tested until hospitalized. The higher proportion of women who are tested, with or without symptoms, is likely a result of this type of selection bias. It is therefore important to note that these results reflect the positivity rates in a community testing program, primarily in adults.

Based on previously reported studies and what we know about the course of disease, these data should not be applied to the hospital setting. Furthermore, new or worsening shortness of breath may be a sign of severity of disease or the presence of a different disease and warrants consultation with a physician even though it may not be predictive of COVID-19 positivity. Nonetheless, our data suggest that public education could be improved around early symptoms vs later stage symptoms. Shortness of breath, in particular, has remained one of the three most heavily emphasized symptoms in public health messaging, while recent loss of taste or smell has received less attention. Our data suggest a re-ordering of symptom emphasis, considering excluding shortness of breath and GI symptoms entirely, could be valuable and make it possible to more accurately screen for the disease.

**Table 2:**
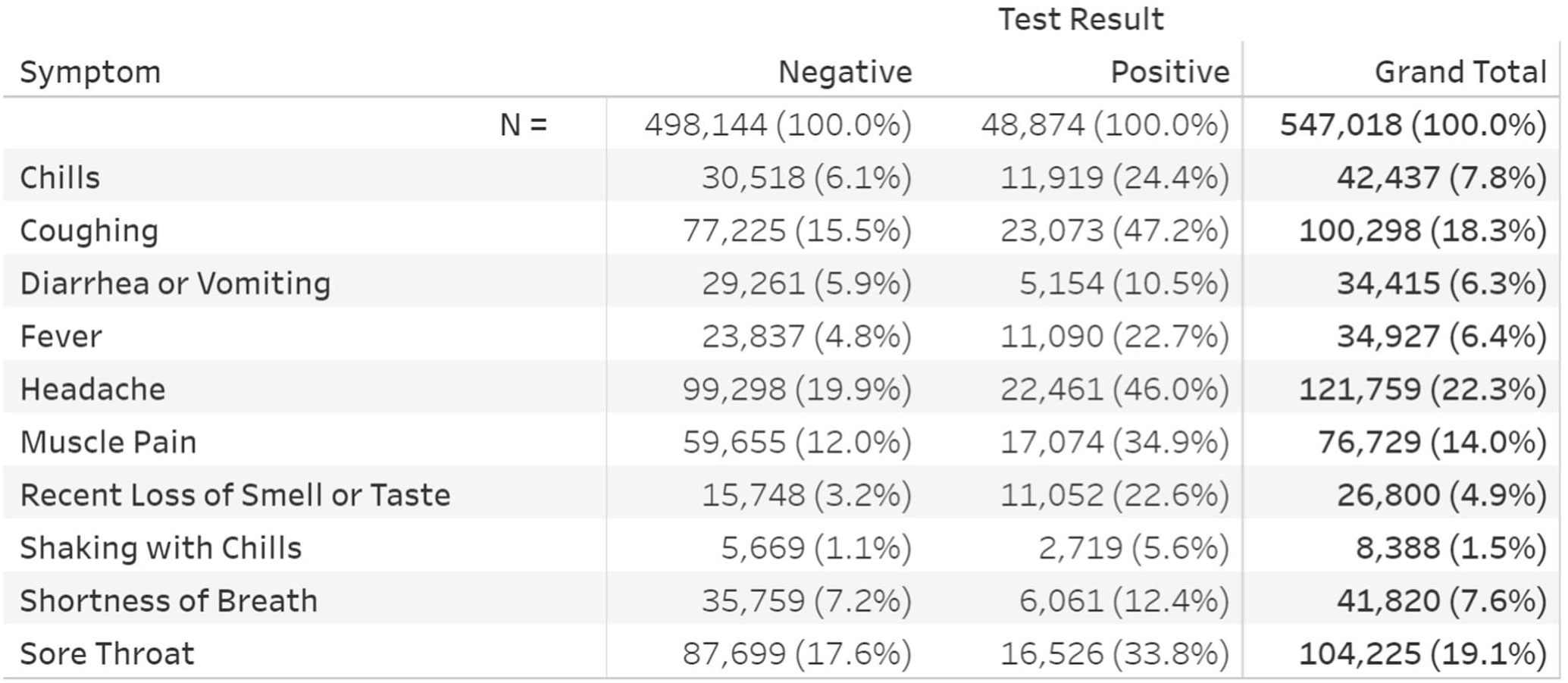
Symptom summary

## Data Availability

Data are only available in aggregate form, as presented in the paper. Due to the presence of protected health information (PHI), which is critical for meaningful analysis, in the form of test dates and county-level geographic identifiers, it is not possible to deidentify or anonymize the data to be shared beyond the presentation of aggregate data.

